# Impact of an Artificial Intelligence Algorithm on Diabetic Retinopathy Grading by Ophthalmology Residents

**DOI:** 10.1101/2023.08.05.23293692

**Authors:** Samantha K. Paul, Christian U. Kim, David Shieh, Xiao Yi Zhou, Ian Pan, Ankur. A Mehra, Warren M. Sobol

**Affiliations:** Department of Ophthalmology, University Hospitals Cleveland Medical Center, Case Western Reserve University School of Medicine, Cleveland, OH; Department of Ophthalmology, Duke University, Durham, NC; Department of Radiology, Brigham and Women’s Hospital, Harvard Medical School, Boston, MA

**Keywords:** diabetic retinopathy, artificial intelligence, deep learning, graduate medical education

## Abstract

**Purpose:** To determine whether AI significantly affects the performance of diabetic retinopathy (DR) grading by ophthalmology residents. Secondary objectives included evaluation of AI’s effects on intergrader variability, self-reported confidence, and decision making.

**Methods:** Four ophthalmology residents at a single academic medical center across all years of training (PGY-2 to PGY-4) analyzed 265 retinal fundus photographs for diabetic retinopathy from a publicly available dataset without and with the assistance of an AI algorithm, separated by a 3-week washout period

**Results:** Overall, there was no significant difference without versus with AI in five-class grading, as measured by QWK, with differences ranging from +0.010-0.017, p=0.09-0.32. No significant difference without and with AI was observed for binary classification of referable DR, except for the specificity of the PGY-3 resident (71.8% to 80%, p=0.019). Intergrader agreement among residents significantly increased with AI (FK +0.072, p=0.0003). Self-reported confidence also significantly increased for 3 out of 4 residents.

**Conclusion:** The use of an AI algorithm did not significantly affect the DR grading performance of ophthalmology residents but did increase intergrader agreement and self-reported confidence. Introducing AI into the ophthalmology residency curriculum may be beneficial as the technology becomes more prevalent.

**Summary Statement:** A cross-sectional study that evaluated the performance of ophthalmology residents grading diabetic retinopathy fundus photographs with and without the assistance of an artificial intelligence algorithm.

## Introduction

Diabetic retinopathy (DR) is a leading cause of vision loss among adults. The prevalence of diabetes continues to increase; by 2045, nearly 700 million people are projected to have diabetes.^1^ Up to 35% of patients with diabetes will have some degree of diabetic retinopathy and nearly 10% will have a visually-threatening complication, such as proliferative diabetic retinopathy (PDR) or diabetic macular edema (DME).^2^ DR is one of the most common pathologies treated by retina specialists, and determining the proper treatment and follow-up relies on accurately assessing the severity, or grade, of DR. Accurate and consistent grading is also important in clinical trials that investigate anatomic and visual acuity outcomes in different stages of DR. The most commonly used grading scale for DR is the International Clinical Disease Severity Scale for Diabetic Retinopathy (ICDR),^3^ which includes 5 stages: 1) no retinopathy, 2) mild non-proliferative diabetic retinopathy (NPDR), 3) moderate NPDR, 4) severe NPDR, and 5) PDR. Despite clear criteria that define each stage of DR, there is substantial intergrader variability among ophthalmologists, and even retina specialists.^4–6^

Many artificial intelligence (AI) algorithms for DR grading have been developed and demonstrate performance comparable or even superior to retina specialists.^7–11^ The vast majority of studies evaluate standalone performance of AI relative to human graders. However, many future implementations of AI in medicine will likely involve augmenting rather than replacing clinicians, and thus studies investigating the impact of AI on human decision making will be important for promoting the adoption of AI.^12^ In ophthalmology, and specifically with regards to DR grading, Sayres et al. studied the impact of an AI algorithm on DR grading by 10 ophthalmologists, demonstrating increased performance and self-reported confidence.^13^

As AI increases its prevalence in ophthalmology, it is important to consider the effects of its adoption not only on experienced clinicians but also on ophthalmologists-in-training. Studies analyzing the impact of AI on residents will be important for determining how best to incorporate AI into resident education. In this study, we studied the impact of AI assistance on the performance and intergrader variability of DR grading by ophthalmology residents.

## Methods

This study was deemed exempt by the institutional review board (IRB) committee of University Hospitals Cleveland Medical Center in Cleveland, Ohio. A sample of 300 fundus photographs from the publicly available Asia Pacific Tele-Ophthalmology Society (APTOS) 2019 Blindness Detection Challenge dataset were used for analysis.^14^ All images in the dataset were already graded according to ICDR criteria by an ophthalmologist for the purposes of the challenge. Thirty-five photographs were excluded due to poor quality or presence of panretinal photocoagulation (PRP) scars. The remaining 265 photographs were then independently re-graded according to ICDR criteria by 2 retina specialists. The reference standard was determined by the median of the 3 grades.

The fundus photographs were then independently graded according to ICDR criteria by 4 ophthalmology residents at different stages of training (2 PGY-2, 1 PGY-3, 1 PGY-4). Each resident provided a DR grade and self-reported confidence score based on a 5-point Likert scale.^15^

The AI algorithm used in this study is an EfficientNet-type convolutional neural network ensemble, trained on the public EyePACS database^16^ and subsequently fine-tuned on 1,000 fundus photographs from the APTOS dataset. No overlap exists between the sample used for fine-tuning and the 265 photographs used for analysis. A demo of this algorithm is available at [redacted]. The software code used to develop the algorithm is available at [redacted].

After a 3-week washout period, the fundus photographs were re-presented to the same 4 residents. The AI algorithm was used to generate predictions for each image, which were now available to the residents. The predictions were presented in a probability-based format, as opposed to a singular class, to allow the residents to see how confident the model was in each class. New DR grades and interpretation confidence scores were provided for each image.

Accuracy and quadratic-weighted Cohen’s kappa (QWK) were used to measure 5-class DR grading performance. QWK is a measure of inter-reader agreement closely related to the traditional (linear) Cohen’s kappa, which accounts for the magnitude of the differences between grades, with a stronger penalty for pairs of grades that are more disparate (e.g., mild versus severe DR is penalized more heavily than mild versus moderate DR). Specificity and sensitivity of referable DR (moderate NPDR, severe NPDR, or proliferative DR) classification was also analyzed. Fleiss’ kappa (FK) was used to quantify groupwise intergrader agreement. Statistical analysis was performed using the Python 3.9 programming language. P-values were calculated using nonparametric permutation tests,^17^ and 95% confidence intervals were calculated using the bootstrap method.^18^ The threshold for statistical significance was set at p=0.05; reported p-values are not corrected for multiple comparisons.

## Results

Based on the reference standard, the dataset comprised 28.3% non-DR cases, 13.2% mild NPDR cases, 27.5% moderate NPDR cases, 21.5% severe NPDR cases, and 9.4% proliferative DR cases. Residents without AI performed similarly for five-class grading, with QWK ranging from 0.866 to 0.906 and accuracy ranging from 65.7% to 71.7% (p>0.05 for all pairwise comparisons). No strong correlation was observed between years of training and QWK (Kendall’s tau=0.18, p=0.72) or accuracy (Kendall’s tau=0.55, p=0.28). Three out of 4 residents classified referable DR with higher sensitivity (96.1% to 98.7%) than specificity (69.1% to 85.5%). One resident (PGY-2_1) classified referable DR with 79.4% sensitivity and 95.5% specificity. The AI model achieved QWK 0.872 and accuracy 73.2% for 5-class grading and 92.9% sensitivity and 92.7% specificity for referable DR. The AI algorithm was 98.7% and 63.2% accurate for images without DR (n=75) and images with mild or worse DR (n=190), respectively, similar to prior work.^13^

### Grading Performance

The results of resident performance for five-class grading and binary classification with and without AI are shown in Figure 1. Residents with AI achieved QWK ranging from 0.881 to 0.923 and accuracy ranging from 68.7% to 77.7%. QWK values were slightly higher with AI (+0.010-0.017), though differences were not statistically significant (p=0.20-0.74). The PGY-3 resident demonstrated a statistically significant improvement in accuracy with AI (+6.0%, p=0.045). The two PGY-2 residents demonstrated a trend towards improvement in accuracy (+5.7%, p=0.06; +3.0%, p=0.32). The PGY-4 resident experienced a minimal drop in accuracy, though this difference was not statistically significant (-0.8%, p=0.88).

**Figure 1.**
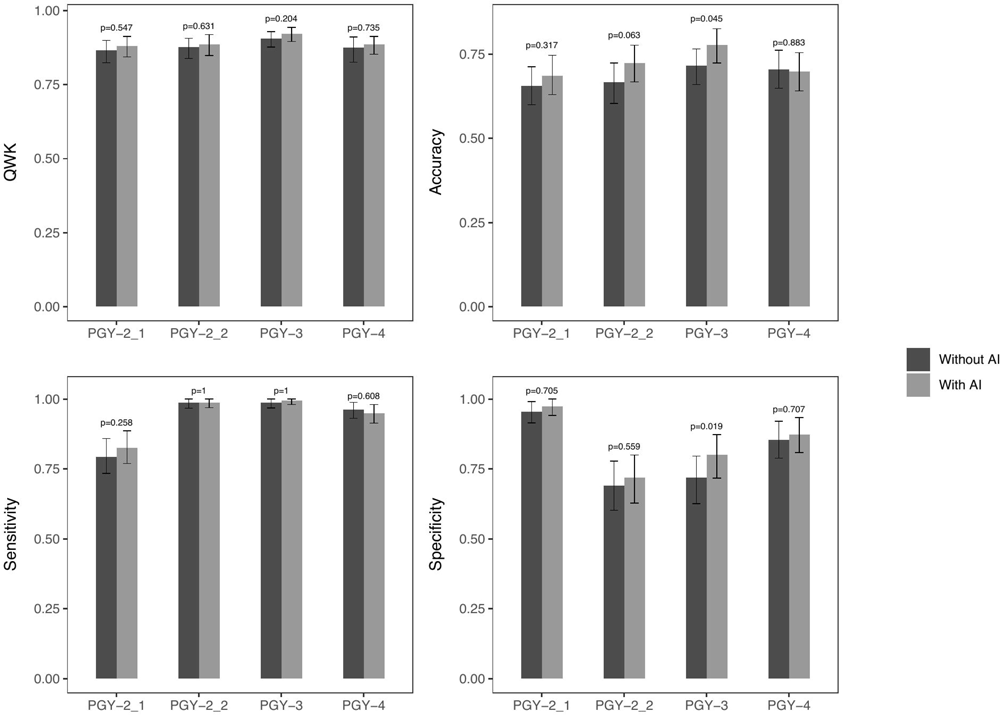
Visual comparison of performance metrics with and without AI across all ophthalmology residents. Error bars represent 95% confidence intervals.

Overall, there was a trend towards improvement with AI versus without in sensitivity and specificity of referable DR classification. The PGY-3 resident demonstrated a statistically significant increase in specificity with AI (71.8% to 80.0%, p=0.019) with similar sensitivity (98.7% to 99.4%, p=1.0). The PGY-4 resident experienced a slight decrease in sensitivity with AI (96.1% to 94.8%, p=0.608). The PGY-2 residents did not demonstrate statistically significant differences in sensitivity (+0.0-3.2%, p=0.258-1.0) or specificity (+1.8-2.7%, p=0.559-0.705).

### Intergrader Agreement

Pairwise intergrader agreement among all graders, as measured by QWK, ranged from 0.81 to 0.91 when the residents did not use AI and 0.83 to 0.92 when the residents used AI (Figure 2). Groupwise intergrader agreement among residents was fair and did not differ significantly from groupwise agreement among experts (FK 0.549 vs. 0.557, p=0.79). AI assistance significantly increased groupwise intergrader agreement among residents (FK 0.549 to 0.621, p=0.003). Intergrader agreement for referable DR was also similar among residents versus experts (FK 0.707 vs 0.724, p=0.55) and increased with AI, though this difference was not statistically significant (FK 0.707 to 0.748, p=0.17).

**Figure 2.**
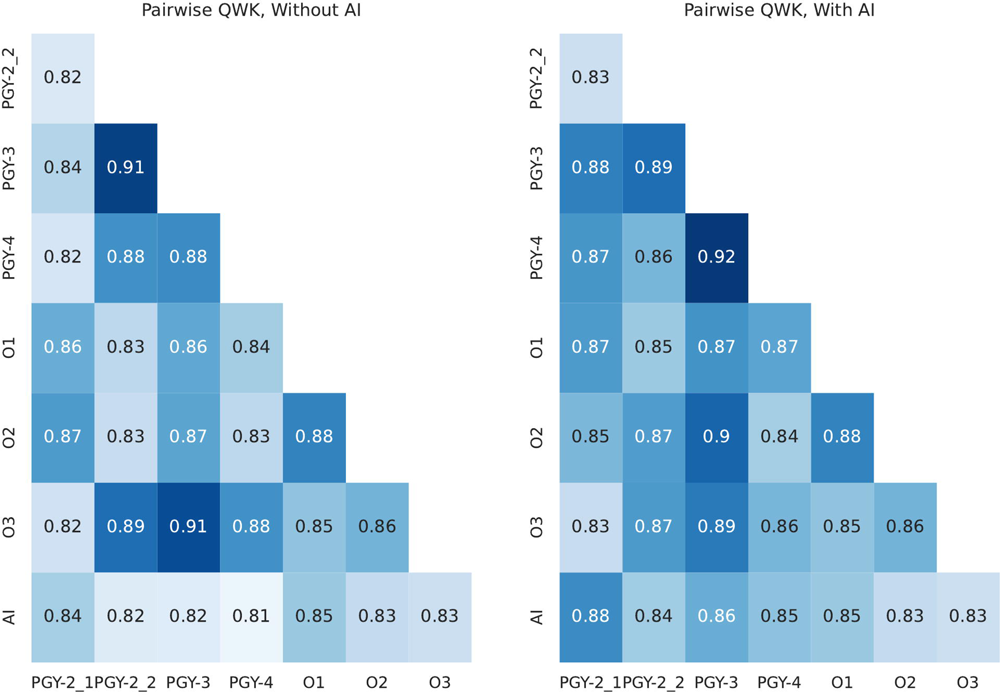
Pairwise quadratic-weighted Cohen’s kappa (QWK) across all graders in the study, without and with AI. Note that only ophthalmology residents used AI assistance; expert grader ratings were the same in both charts.

### Self-Reported Confidence

Three out of 4 residents experienced significant increases in self-reported confidence with AI (Figure 4), ranging from +0.32 to +0.60. The PGY-4 resident did not experience a statistically significant change in confidence (-0.06, p=0.458). The mean confidence scores reported when the residents used AI were significantly lower for changed cases (i.e., cases for which the grade with AI differed from the initial grade without AI) compared to unchanged cases (-0.13 to -0.60, p<0.001 to p=0.027). Confidence was higher for correct versus incorrect cases, both without AI (0.37 to 0.78, p<0.001 to p=0.005), and with AI (0.12 to 0.73, p<0.001 to p=0.065).

### Decision Making

We define resident-AI agreement as cases for which the resident’s prediction with AI was concordant with the standalone AI prediction during the second stage of labeling (i.e., the resident’s final prediction with AI agreed with the AI prediction). Agreement ranged from 72.5% to 78.9%. QWK against the reference standard for concordant cases was significantly higher than for discordant cases (0.920-0.956 vs. 0.632-0.731, p<0.001) across all residents (Figure 3). This difference persisted when excluding cases without DR, though differences were no longer statistically significant (0.691-0.821 vs. 0.565-0.722, p=0.08-0.68).

**Figure 3.**
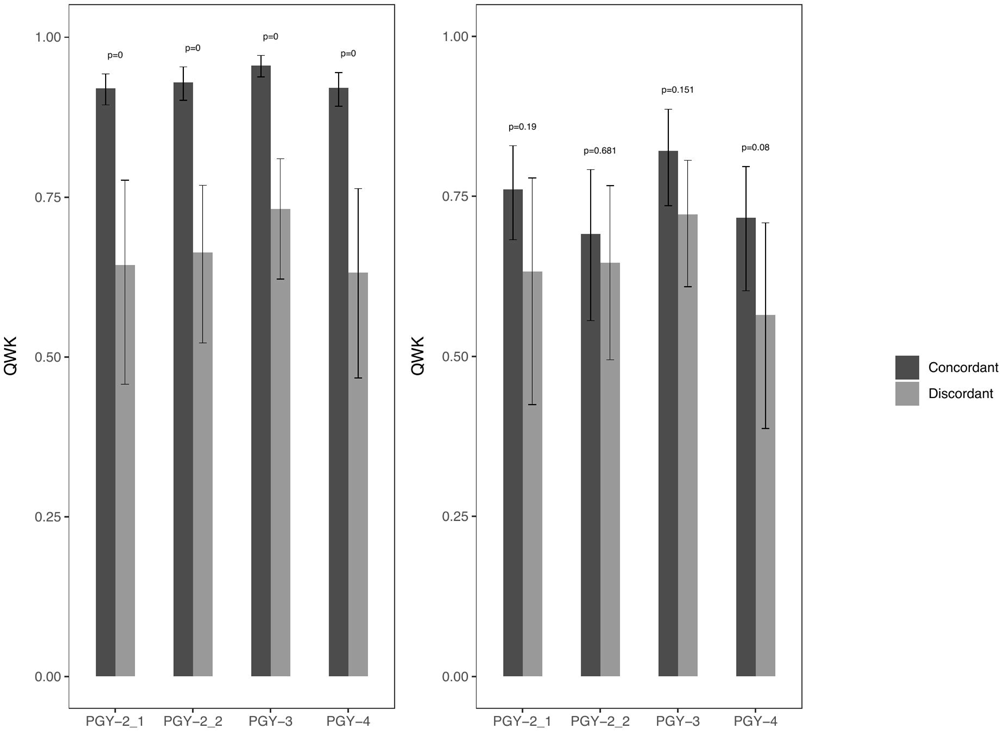
Comparison of quadratic-weighted Cohen’s kappa (QWK) for concordant (agreement between AI and final resident prediction with AI) and discordant (disagreement between AI and final resident prediction with AI) for all cases (left) and cases excluding those without diabetic retinopathy (right). Error bars demonstrate 95% confidence intervals.

For discordant cases, nearly all differences were by 1 grade (71.2-86.3%) or 2 grades (16.1-27.1%). Among these cases, for 3 out of 4 residents, the most common behavior was to grade 1 higher than the AI; one PGY-2 resident most frequently graded 1 less than the AI. Figure 4a illustrates the distribution of differences for discordant cases. One common case resulted in a grade difference from the AI of 3 for all residents; this case was graded proliferative DR by all residents and experts, while the AI graded it as mild. Upon further review, this was a case of inactive PDR.

**Figure 4.**
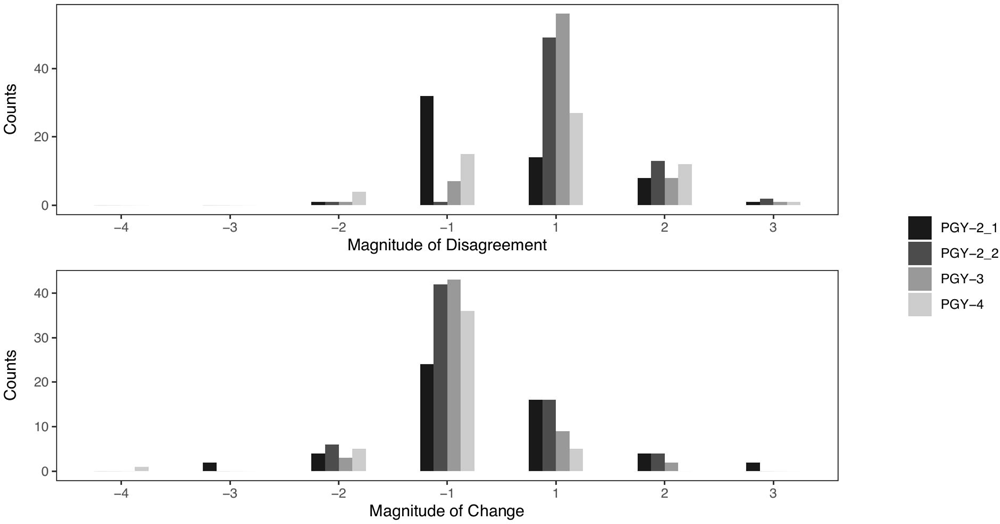
Distribution of differences (resident with AI minus AI) in grade between the AI and final resident predictions with AI for discordant cases (top). Distribution of changes (resident with AI minus resident without AI) in grade made by the resident with AI versus without AI for changed cases (bottom).

For 3 out of 4 residents, the majority of changed predictions were from incorrect to correct grades (51.9% to 63.2%). For the PGY-4 resident, 48.9% of changed predictions were from correct to incorrect and 44.7% were from correct to incorrect. When analyzing the magnitude and direction of change, the most common type of change across all residents was to downgrade by a single grade (Figure 4b). This complements the observation that the mean AI grade over all cases was slightly lower (2.49) than the residents’ mean grades without AI (2.51 to 2.89).

**Table 1.** Summary of performance metrics and self-reported confidence with and without AI for across all ophthalmology residents.

|  | Without AI | With AI | Difference | p-value |
| --- | --- | --- | --- | --- |
| **QWK** |  |  |  |  |
| PGY-2_1 | 0.866 (0.824, 0.899) | 0.881 (0.844, 0.913) | 0.015 (-0.026, 0.063) | 0.547 |
| PGY-2_2 | 0.877 (0.839, 0.907) | 0.887 (0.849, 0.919) | 0.010 (-0.029, 0.049) | 0.631 |
| PGY-3 | 0.906 (0.878, 0.930) | 0.923 (0.896, 0.944) | 0.017 (-0.009, 0.044) | 0.204 |
| PGY-4 | 0.876 (0.826, 0.912) | 0.886 (0.853, 0.913) | 0.011 (-0.021, 0.053) | 0.735 |
| **Accuracy** |  |  |  |  |
| PGY-2_1 | 65.7% (60.0%, 71.3%) | 68.7% (63.0%, 74.7%) | 3.0% (-1.9%, 8.3%) | 0.317 |
| PGY-2_2 | 66.8% (60.4%, 72.5%) | 72.5% (66.8%, 77.7%) | 5.7% (-0.4%, 11.7%) | 0.063 |
| PGY-3 | 71.7% (66.0%, 76.6%) | 77.7% (72.2%, 82.6%) | 6.0% (0.4%, 11.3%) | 0.045 |
| PGY-4 | 70.6% (64.9%, 76.2%) | 69.8% (64.2%, 75.5%) | -0.8% (-5.7%, 3.8%) | 0.883 |
| **Sensitivity** |  |  |  |  |
| PGY-2_1 | 79.4% (73.5%, 85.8%) | 82.6% (76.9%, 88.7%) | 3.2% (-1.3%, 8.0%) | 0.258 |
| PGY-2_2 | 98.7% (96.7%, 100%) | 98.7% (96.9%, 100%) | 0.0% (-2.5%, 2.5%) | 1 |
| PGY-3 | 98.7% (96.8%, 100%) | 99.4% (98.0%, 100%) | 0.6% (0.0%, 2.1%) | 1 |
| PGY-4 | 96.1% (93.1%, 98.8%) | 94.8% (91.4%, 98.0%) | -1.3% (-3.8%, 1.3%) | 0.608 |
| **Specificity** |  |  |  |  |
| PGY-2_1 | 95.5% (91.4%, 99.0%) | 97.3% (94.1%, 100%) | 1.8% (-2.7%, 6.4%) | 0.705 |
| PGY-2_2 | 69.1% (60.2%, 77.8%) | 71.8% (62.7%, 80.0%) | 2.7% (-3.3%, 8.3%) | 0.559 |
| PGY-3 | 71.8% (62.6%, 79.6%) | 80.0% (71.7%, 87.3%) | 8.2% (2.5%, 14.4%) | 0.019 |
| PGY-4 | 85.5% (78.8%, 92.0%) | 87.3% (80.8%, 93.3%) | 1.8% (-2.8%, 6.4%) | 0.707 |
| **Confidence** |  |  |  |  |
| PGY-2_1 | 4.25 (4.16, 4.35) | 4.86 (4.81, 4.90) | 0.60 (0.51, 0.69) | <0.0001 |
| PGY-2_2 | 3.63 (3.49, 3.77) | 4.03 (3.91, 4.15) | 0.40 (0.26, 0.55) | <0.0001 |
| PGY-3 | 4.20 (4.12, 4.28) | 4.52 (4.45, 4.59) | 0.32 (0.23, 0.42) | <0.0001 |
| PGY-4 | 3.69 (3.57, 3.80) | 3.62 (3.52, 3.73) | -0.06 (-0.16, 0.04) | 0.46 |

## Discussion

In this work, we examined the impact of AI on DR grading by ophthalmology residents. Our goal was to study the impact of AI at the trainee level, as most prior work has focused on experienced ophthalmologists. Han et al. studied the use of an AI algorithm as a tool for training junior ophthalmology residents and medical students in DR grading,^19^ whereas our work studied the influence of AI on resident performance, self-reported confidence, and intergrader variability. We also used publicly available datasets and open sourced our code and AI algorithms to encourage reproducibility and further research in this area.

Our results found no statistically significant difference between resident performance with and without AI across a variety of metrics, except one PGY-3 resident who had a statistically significant improvement in accuracy and specificity. These results are in contrast to the previous study performed by Sayres et al., which primarily focused on attending ophthalmologists.^13^ Potential explanations include a lower sample size used for analysis, different distribution of DR grades, differences in clinical experience, and differences in the AI algorithm.

Though performance did not improve with AI, there was a statistically significant improvement in intergrader agreement associated with AI. This is not unexpected; as all residents were provided with the same AI predictions, as long as the AI somewhat influenced their final grade, the grades would converge towards the AI predictions and reduce variability. Improving intergrader agreement can improve standardization of care and may particularly be useful for clinical trials that depend on these grades. AI also significantly increased self-reported confidence in 3 out of 4 residents, suggesting its potential use in aiding less experienced ophthalmologists as a second reader.

Overall, agreement was fairly high between the standalone AI prediction and the residents’ final grades with AI (72.5-78.8%). Higher performance was achieved for cases where the resident and AI agreed on the final prediction compared to cases where there was disagreement. Thus, agreement between the resident and AI can serve as a strong indicator that the determined grade is accurate. Conversely, resident and AI disagreement can be used to select challenging cases for additional attending review, giving rise to more learning opportunities. When analyzing changed predictions (i.e., where the resident grade with AI differed from the resident grade without AI), we found that for 3 out of 4 residents, the majority of changed predictions resulted in a change from incorrect to correct. The most common type of change across all residents was to downgrade a study by 1 grade.

Several limitations exist in our study. The AI predictions were not accompanied by any method of explainability, which creates challenges for incorporating the predictions into one’s decision making process, especially for cases where there is disagreement. However, previous work demonstrated that including saliency maps with the AI prediction, one of the most common methods of explainability, did not improve performance. Another potential limitation was that our distribution of diabetic retinography grades was oversampled to include more positive cases to ensure enough variability among DR grades in the dataset given the relatively low sample size. However, this is not necessarily representative of the expected real-world distribution, where most cases would be negative for DR. Finally, our study only included residents from a single institution and may not reflect the behavior of residents at other training programs. Future work including a multi-institutional cohort of residents is needed to better understand the nuances of the impact of AI on ophthalmology residents.

## Conclusions

Overall, performance of DR grading by ophthalmology residents was not significantly affected by the use of an AI algorithm, though a trend towards improvement was observed. However, use of the AI algorithm improved intergrader agreement and self-reported confidence. In addition, cases where the resident and AI agreed upon the prediction were more likely to be correct than cases where there was disagreement, suggesting that, at the trainee level, AI may be most useful as an adjunct in determining when an assessment would most benefit from a higher level of expertise.

## Data Availability

All data produced in the present study are available upon reasonable request to the authors

## References

1. Saeedi P, Petersohn I, Salpea P, et al. Global and regional diabetes prevalence estimates for 2019 and projections for 2030 and 2045: Results from the International Diabetes Federation Diabetes Atlas, 9th edition. Diabetes Res Clin Pract. 2019;157:107843. doi:10.1016/j.diabres.2019.107843

2. Yau JW, Rogers SL, Kawasaki R, Lamoureux EL, Kowalski JW, Bek T, Chen SJ, Dekker JM, Fletcher A, Grauslund J, Haffner S, Hamman RF, Ikram MK, Kayama T, Klein BE, Klein R, Krishnaiah S, Mayurasakorn K, O’Hare JP, Orchard TJ, Porta M, Rema M, Roy MS, Sharma T, Shaw J, Taylor H, Tielsch JM, Varma R, Wang JJ, Wang N, West S, Xu L, Yasuda M, Zhang X, Mitchell P, Wong TY; Meta-Analysis for Eye Disease (META-EYE) Study Group. Global prevalence and major risk factors of diabetic retinopathy. Diabetes Care. 2012 Mar;35(3):556–64. doi: 10.2337/dc11-1909. Epub 2012 Feb 1. PMID: 22301125; PMCID: PMC3322721.

3. Wilkinson CP, Ferris FL 3rd, Klein RE, et al. Proposed international clinical diabetic retinopathy and diabetic macular edema disease severity scales. Ophthalmology. 2003;110(9):1677–1682. doi:10.1016/S0161-6420(03)00475-5

4. Liu Y, Rajamanickam VP, Parikh RS, Loomis SJ, Kloek CE, Kim LA, Hitchmoth DL, Song BJ, Xerras DC, Pasquale LR. Diabetic Retinopathy Assessment Variability Among Eye Care Providers in an Urban Teleophthalmology Program. Telemed J E Health. 2019 Apr;25(4):301–308. doi: 10.1089/tmj.2018.0019.

5. Krause J, Gulshan V, Rahimy E, et al. Grader Variability and the Importance of Reference Standards for Evaluating Machine Learning Models for Diabetic Retinopathy. Ophthalmology. 2018;125(8):1264–1272. doi:10.1016/j.ophtha.2018.01.034

6. Grzybowski A, Brona P, Krzywicki T, Gaca-Wysocka M, Berlińska A, Święch A. Variability of Grading DR Screening Images among Non-Trained Retina Specialists. J Clin Med. 2022 May 31;11(11):3125. doi: 10.3390/jcm11113125. PMID: 35683522; PMCID: PMC9180965.

7. Ting DSW, Cheung CY, Lim G, et al. Development and Validation of a Deep Learning System for Diabetic Retinopathy and Related Eye Diseases Using Retinal Images From Multiethnic Populations With Diabetes. JAMA. 2017;318(22):2211–2223. doi:10.1001/jama.2017.18152

8. Gulshan V, Peng L, Coram M, et al. Development and Validation of a Deep Learning Algorithm for Detection of Diabetic Retinopathy in Retinal Fundus Photographs. JAMA. 2016;316(22):2402–2410. doi:10.1001/jama.2016.17216

9. Dai L, Wu L, Li H, et al. A deep learning system for detecting diabetic retinopathy across the disease spectrum. Nat Commun. 2021;12(1):3242. Published 2021 May 28. doi:10.1038/s41467-021-23458-5

10. Gargeya R, Leng T. Automated Identification of Diabetic Retinopathy Using Deep Learning. Ophthalmology. 2017;124(7):962–969. doi:10.1016/j.ophtha.2017.02.008

11. Lee AY, Yanagihara RT, Lee CS, et al. Multicenter, Head-to-Head, Real-World Validation Study of Seven Automated Artificial Intelligence Diabetic Retinopathy Screening Systems. Diabetes Care. 2021;44(5):1168–1175. doi:10.2337/dc20-1877

12. Langlotz CP. Will Artificial Intelligence Replace Radiologists? Radiology: Artificial Intelligence. 2019;1(3):e190058.

13. Sayres R, Taly A, Rahimy E, et al. Using a Deep Learning Algorithm and Integrated Gradients Explanation to Assist Grading for Diabetic Retinopathy. Ophthalmology. 2019;126(4):552–564.

14. APTOS 2019 Blindness Detection. https://www.kaggle.com/c/aptos2019-blindness-detection. Accessed April 9, 2022.

15. Likert, R. A technique for the measurement of attitudes. Archives of Psychology. 1932;22(140): 55.

16. EyePACS. http://www.eyepacs.com/data-analysis. Accessed April 9, 2022.

17. Ojala M and Garriga GC. Permutation Tests for Studying Classifier Performance. 2009 Ninth IEEE International Conference on Data Mining, 2009:908-913.

18. Efron, B. Bootstrap Methods: Another Look at the Jackknife. The Annals of Statistics. 1979;7(1):1–26.

19. Han R, Yu W, Chen H, Chen Y. Using artificial intelligence reading label system in diabetic retinopathy grading training of junior ophthalmology residents and medical students. BMC Med Educ. 2022 Apr 9;22(1):258. doi: 10.1186/s12909-022-03272-3.

